# University students and staff able to maintain low daily contact numbers during various COVID-19 guideline periods

**DOI:** 10.1101/2021.01.19.21250097

**Authors:** Adam Trickey, Emily Nixon, Hannah Christensen, Adam Finn, Amy Thomas, Caroline Relton, Clara Montgomery, Gibran Hemani, Jane Metz, Josephine G. Walker, Katy Turner, Rachel Kwiatkowska, Sarah Sauchelli, Leon Danon, Ellen Brooks-Pollock

**Affiliations:** Population Health Sciences, University of Bristol, Bristol, UK; School of Biological Sciences, University of Bristol, Bristol, UK; NIHR Health Protection Research Unit in Behavioural Science and Evaluation at University of Bristol, Bristol, UK; School of Cellular and Molecular Medicine, University of Bristol, Bristol, UK; Bristol Veterinary School, University of Bristol, Bristol, UK; Bristol Children’s Vaccine Centre, University of Bristol, Bristol, United Kingdom; National Institute for Health Research Bristol Biomedical Research Centre, University Hospitals of Bristol and Weston NHS Foundation Trust and University of Bristol; Computer Science, University of Exeter, Exeter, UK; Alan Turing Institute, British Library, London, UK

## Abstract

**Introduction:** UK universities re-opened in September 2020, despite the on-going coronavirus epidemic. During the first term, various national social distancing measures were introduced, including banning groups of >6 people and the second lockdown in November. COVID-19 can spread rapidly in university-settings, and students’ adherence to social distancing measures is critical for controlling transmission.

**Methods:** We measured university staff and student contact patterns via an online, longitudinal survey capturing self-reported contacts on the previous day. We investigated the change in contacts associated with COVID-19 guidance periods: post-first lockdown (23/06/2020-03/07/2020), relaxed guidance period (04/07/2020-13/09/2020), “rule-of-six” period (14/09/2020-04/11/2020), and the second lockdown (05/11/2020-25/11/2020).

**Results:** 722 staff (4199 responses) (mean household size: 2.6) and 738 students (1906 responses) (mean household size: 4.5) were included in the study. Contact number decreased with age. Staff in single-person households reported fewer contacts than individuals in 2-and 3-person households, and individuals in 4-and 5-person households reported more contacts.

For staff, daily contacts were higher in the relaxed guidance and “rule-of-six” periods (means: 3.2 and 3.5, respectively; medians: 3) than the post-first lockdown and second lockdown periods (means: 4.5 and 5.4, respectively; medians: 2). Few students responded until 05/10/2020, after which the median student contacts was 2 and the mean was 5.7, until the second lockdown when it dropped to 3.1.

**Discussion:** University staff and students responded to national guidance by altering their social contacts. The response in staff and students was similar, suggesting that students are able to adhere to social distancing guidance while at university.

## Background

Due to the COVID-19 pandemic, different countries implemented different laws in 2020 to limit people’s contacts and therefore COVID-19 transmission^1^. In the UK, the first lockdown implemented on 23/03/2020, legally restricted movement of people from their place of residence, with movement only being permittable when seeking healthcare, to exercise (alone/with household members), to purchase necessities, or to assist vulnerable persons. Subsequently the laws were eased from 01/06/2020^2^. However, on 14/09/2020 the guidance was again tightened and then England entered a second lockdown on 05/11/2020, which involved shutting down non-essential shops, working from home where possible, restricting gatherings to two people meeting outside in a public place, but with schools and universities remaining open^3^. Before the second lockdown, some UK areas had restrictions tightened above the national guidance due to higher transmission rates through the implementation of tiers or legislation in devolved nations, but Bristol and the Southwest of England remained in the lowest tier throughout this period due to the low overall COVID-19 transmission rate^4^.

The first lockdown forced universities to move teaching online^5^, including the University of Bristol (UoB). Universities began the 2020/21 term in the autumn, when reported daily COVID-19 cases were rising nationally^6^. Students migrated from around the UK and abroad to attend the new term. Although, university students are mostly young and are therefore less likely to be severely affected by COVID-19 morbidity and mortality than other groups, some may still be medically vulnerable^7^. Meanwhile, university staff are more representative of the working-age general population and tend to be older and are therefore more likely to be affected by COVID-19 morbidity and mortality.

For UoB’s 2020/21 term, students returned towards the end of September. The UoB adopted a “blended” teaching approach, including a mixture of face-to-face and online teaching. Students living in university halls of residence were divided into households (“living circles”) and were instructed not to host non-residents in their flat but government social distancing guidelines applied outside the flat^8^. Students that test positive are required to isolate along with their household^8^.

Despite COVID-19 restrictions, outbreaks of COVID-19 occurred across many UK universities during autumn 2020^9^. For UoB, there were outbreaks among students but few cases amongst staff: UoB reported 1722 positive tests among students from 14/10/2020-01/11/2020, roughly 7% of students, compared with 48 positive tests among staff (<1%)^8^. Hundreds of students (mostly undergraduates) in university-owned halls of residence were told to self-isolate during the beginning of term.

There is little evidence to quantify the effect that the various COVID-19 restrictions in the UK have had on the number of contacts of individuals: a key driver of COVID-19 transmission. On 23/06/2020, we launched an online survey detailing the contacts and behaviours of staff and students at the UoB, with the survey continuing into the autumn term. We aimed to investigate whether there were differences in contact patterns for UoB staff and students between the periods before and during the autumn 2020 COVID-19 lockdown, and to quantify these differences.

## Methods

CONQUEST (COroNavirus QUESTionnaire) is a survey that started on 23/06/2020 asking about contacts, behaviour, and potential SARS-Cov-2 symptoms for staff and students at UoB. Survey participants complete an initial questionnaire including questions on background demographics and then have the option to fill out a shorter, recurring version of the questionnaire on contacts, symptoms, and whether they have had COVID-19. The recurring questionnaire was initially every 14 days and then every 8 days as of 13/09/2020 (see supplement for details). It was not possible to advertise the survey to students at the end of the 2019/2020 academic year via direct email and only light touch promotion was granted for social media. The survey was advertised to staff via email and newsletters during June and July 2020.. Approval was granted for a larger targeting campaign for students when they returned to the university for the 2020/2021 academic year in September. Here, we present the data up to 25/11/2020.

### Survey

Survey data were collected using UoB’s REDCap Electronic Data Capture^10,11^. The initial survey (see supplementary materials) captured demographic information on participants and asked about symptoms in the last 7 days, whether they had sought medical attention for these symptoms, whether they had been self-isolating in the last 7 days, and their COVID-19 status.

Participants were asked about contacts they had had on the previous day, which were split into three types:

1. Individual contacts: those who they spoke to in person one-on-one, including those in their household and support bubble.
2. Other contacts: if they spoke in person to many people one-on-one in the same setting (but they did not have the opportunity to speak to each other), for example, as part of working in a customer service role in a shop.
3. Group contacts: large groups of individuals in the same setting (for example, sports teams, tutorials, lectures, religious services, large gatherings with friends and family).

Further information on the questions asked about each of these contact types is given in the supplementary materials along with the full questionnaire. On 13/09/2020 amendments were made to the questionnaire (see supplement). Data are available from the corresponding author.

### Inclusion criteria

We excluded responses where the survey was incomplete. We only include respondents that live in the Southwest of England as this region (including Bristol) remained in the UK government COVID-19 tier-1 throughout the existence of these tiers for the study period presented here.

### COVID-19 guidance periods

Table 1 presents key COVID-19 guidance implementation dates and dates relating to the CON-QUEST survey. The periods of COVID-19 restrictions were stratified as follows:

**Table 1:** List of key events relating to COVID-19 restrictions and the CON-QUEST survey around the study period^13^

| **Date** | **Event** |
| --- | --- |
| 1<sup>st</sup> June 2020 | **Health Protection (Coronavirus, Restrictions) (England) (Amendment No. 3) Regulations 2020**<br>Spring lockdown ends, with outdoor sports allowed, six person gatherings allowed outside, gatherings prohibited indoors (with exceptions including education).<br>Public transport for non-essential travel is not allowed<br>The University of Bristol 2019/2020 academic teaching year ends. |
| 15<sup>th</sup> June 2020 | **Health Protection (Coronavirus, Restrictions) (England) (Amendment No. 4) Regulations 2020**<br>**Health Protection (Coronavirus, Wearing of Face Coverings on Public Transport) (England) Regulations 2020**<br>One-adult households can be linked with other households for permitted overnight gatherings “support bubbles”<br>General re-opening of English retail shops<br>Year 10 and year 12 secondary school pupils return to school<br>Travelers on public transport must wear a face covering |
| 23<sup>rd</sup> June 2020 | **CON-QUEST survey launched** |
| 4<sup>th</sup> July 2020 | **Health Protection (Coronavirus, Restrictions) (No. 2) (England) Regulations 2020**<br>Physical distancing guidance relaxed to 1 metre. Hospitality venues and hairdressers reopen. Two households can meet indoors. Most indoor gatherings of any size are now allowed. Local areas can be placed into lockdown. |
| 11<sup>th</sup> July 2020 | **Health Protection (Coronavirus, Restrictions) (No. 2) (England) (Amendment) Regulations 2020**<br>Outdoor swimming pools and water parks to re-open |
| 13<sup>th</sup> July 2020 | **Health Protection (Coronavirus, Restrictions) (No. 2) (England) (Amendment) Regulations 2020**<br>The remainder of the amendment comes into effect, with beauty salons (and similar venues) re-opening |
| 18<sup>th</sup> July 2020 | **Health Protection (Coronavirus, Restrictions) (No. 3) (England) Regulations 2020**<br>People can use public transport for non-essential journeys.<br>Local authorities have new powers to close shops and cancel events |
| 24<sup>th</sup> July 2020 | **Health Protection (Coronavirus, Wearing of Face Coverings in a Relevant Place) (England) Regulations 2020**<br>Members of the public must wear a face covering in most indoor shops, banks, and public transport hubs |
| 8<sup>th</sup> August | **Health Protection (Coronavirus, Wearing of Face Coverings in a Relevant Place) (England) (Amendment) Regulations 2020**<br>Face coverings must be worn in places of worship, community centres, public areas of hotels, museums, libraries, and similar locations. |
| 13<sup>th</sup> September 2020 | **Alterations to CON-QUEST survey come into effect in advance of the new COVID-19 regulations and the start of the University of Bristol 2020/2021 academic year** |
| 14<sup>th</sup> September | **Health Protection (Coronavirus, Restrictions) (No. 4) (England) Regulations 2020**<br>Limit to the number of persons in an indoor gathering to no more than 6, with some |
| 2020 | exceptions such as education, work, and organised sports |
| 5th<br>October<br>2020 | The start of a communications campaign to UoB students containing details of the CON-QUEST survey |
| 30<sup>th</sup><br>October<br>2020 | A second lockdown is announced but does not come into effect |
| 5<sup>th</sup><br>November<br>2020 | **A four-week lockdown comes into effect**<br>People must remain at home, including for work. With exceptions for schools, universities, manufacturing businesses, construction sites, healthcare, and supermarkets. |

- Post-first lockdown: Survey start (23/06/2020) to the day before the 2^nd^, more lenient set of COVID-19 regulations were implemented (03/07/2020).
- Relaxed period: 2^nd^ COVID-19 regulations implementation (04/07/2020) to the day before the 4^th^ set of COVID-19 laws were implemented (13/09/2020).
- “Rule-of-six” period: 4^th^ COVID-19 regulations (14/09/2020) to the day before the 2^nd^ lockdown (04/11/2020).
- 2^nd^ Lockdown: 2^nd^ lockdown start (05/11/2020) to data cut-off (25/11/2020).

### Analyses

To make the dataset more representative of UoB’s staff and student populations, weighting was used, described further in the supplementary materials.

We investigated the associations between the overall number of contacts on the previous day with demographics and behaviours using univariable and multivariable negative binomial regression modelling, stratified for staff and students. All variables included in these models are presented in the relevant results tables. Note that cardinal symptoms are defined as loss of taste or smell, fever, persistent cough^12^ and all postgraduates were assigned to the 4+ year group to differentiate them from undergraduates in their first year of study.

### Ethical approval

Ethical approval was granted on the 14/05/2020 by the Health Sciences University Research Ethics Committee at the University of Bristol (ID 104903), with four amendment requests approved on the 22/05/2020, 09/06/2020, 27/08/2020, and 07/09/2020 to update the relevance of the questions or to make the survey faster and easier to complete. All research was performed in accordance with the University of Bristol Ethics of Research Policy and Procedure (http://www.bristol.ac.uk/media-library/sites/red/documents/research-governance/Ethics_Policy_v8_03-07-19.pdf). Participants were aged ≥18, voluntarily opted-in to the study and were required to give their informed consent before starting the survey.

## Results

Included over the entire survey period were 722 staff, with repeat questionnaires leading to 4199 responses, whilst for students there were 738 participants and 1906 questionnaire responses. The median ages of the staff and students were 42 (interquartile range [IQR]: 34-51) and 22 (IQR: 19-25), respectively. The median household size for staff was 2 (IQR: 1-3; mean: 2.6) and 3 for students (IQR: 2-5; mean: 4.5). Most staff participants were recruited between 23/06/2020-13/09/2020 (95.3%), whilst 20.7% of students were recruited between these dates (table 2). Due to the communications campaign, most students (78.0%) were recruited between the 14/09/2020-04/11/2020, whilst 4.3% of staff were recruited during this period. In the weighted analyses there were 1623 staff responses between 14/09/2020-04/11/2020 and 628 from the 05/11/2020-24/11/2020. For the students, these numbers were 1314 and 333, respectively.

**Table 2:**
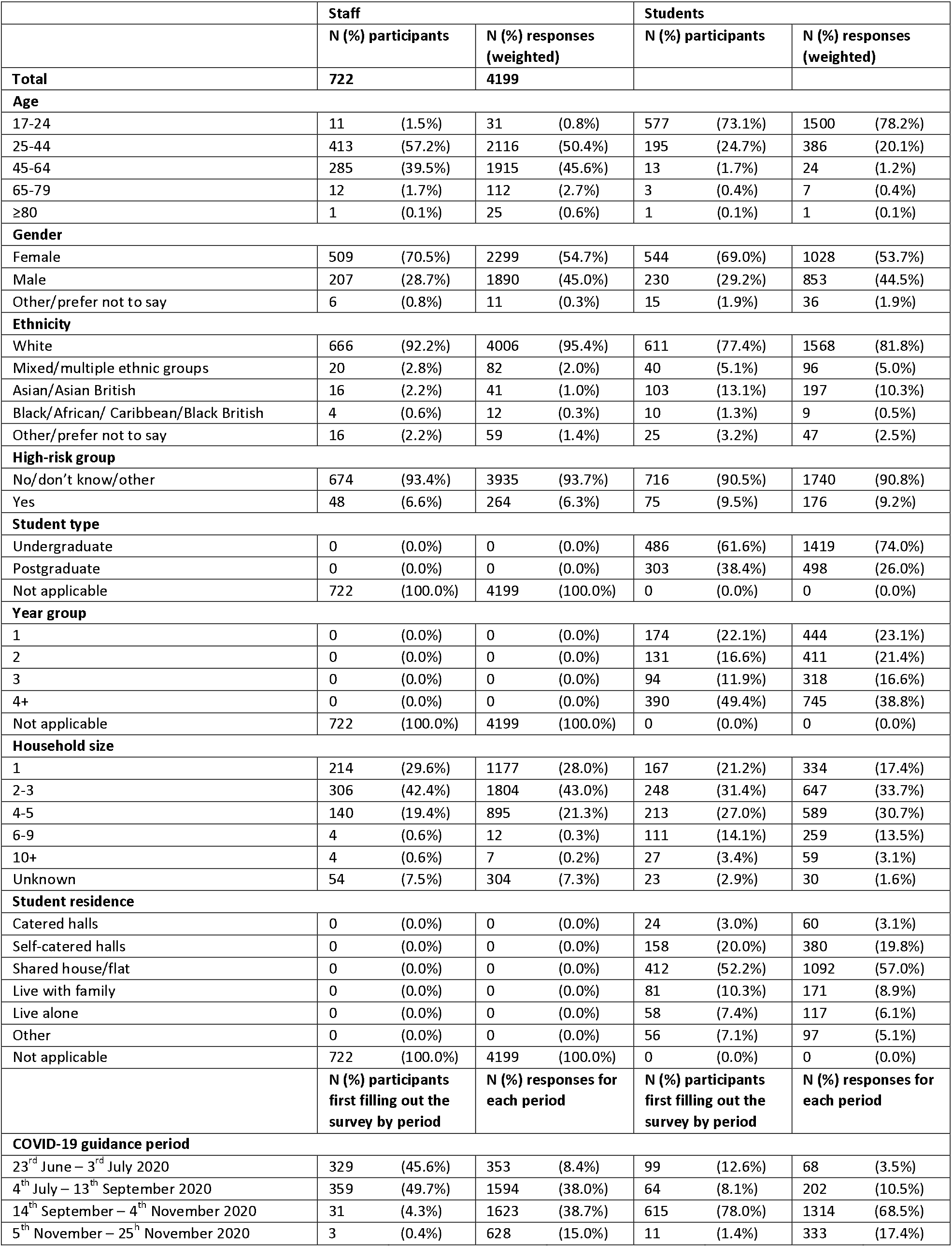
Characteristics of survey participants and responses (weighted)

|  | Staff |  | Students |  |
| --- | --- | --- | --- | --- |
|  | N (%) participants | N (%) responses (weighted) | N (%) participants | N (%) responses (weighted) |
| **Total** | **722** | **4199** |  |  |
| **Age** |  |  |  |  |
| 17-24 | 11 (1.5%) | 31 (0.8%) | 577 (73.1%) | 1500 (78.2%) |
| 25-44 | 413 (57.2%) | 2116 (50.4%) | 195 (24.7%) | 386 (20.1%) |
| 45-64 | 285 (39.5%) | 1915 (45.6%) | 13 (1.7%) | 24 (1.2%) |
| 65-79 | 12 (1.7%) | 112 (2.7%) | 3 (0.4%) | 7 (0.4%) |
| ≥80 | 1 (0.1%) | 25 (0.6%) | 1 (0.1%) | 1 (0.1%) |
| **Gender** |  |  |  |  |
| Female | 509 (70.5%) | 2299 (54.7%) | 544 (69.0%) | 1028 (53.7%) |
| Male | 207 (28.7%) | 1890 (45.0%) | 230 (29.2%) | 853 (44.5%) |
| Other/prefer not to say | 6 (0.8%) | 11 (0.3%) | 15 (1.9%) | 36 (1.9%) |
| **Ethnicity** |  |  |  |  |
| White | 666 (92.2%) | 4006 (95.4%) | 611 (77.4%) | 1568 (81.8%) |
| Mixed/multiple ethnic groups | 20 (2.8%) | 82 (2.0%) | 40 (5.1%) | 96 (5.0%) |
| Asian/Asian British | 16 (2.2%) | 41 (1.0%) | 103 (13.1%) | 197 (10.3%) |
| Black/African/ Caribbean/Black British | 4 (0.6%) | 12 (0.3%) | 10 (1.3%) | 9 (0.5%) |
| Other/prefer not to say | 16 (2.2%) | 59 (1.4%) | 25 (3.2%) | 47 (2.5%) |
| **High-risk group** |  |  |  |  |
| No/don't know/other | 674 (93.4%) | 3935 (93.7%) | 716 (90.5%) | 1740 (90.8%) |
| Yes | 48 (6.6%) | 264 (6.3%) | 75 (9.5%) | 176 (9.2%) |
| **Student type** |  |  |  |  |
| Undergraduate | 0 (0.0%) | 0 (0.0%) | 486 (61.6%) | 1419 (74.0%) |
| Postgraduate | 0 (0.0%) | 0 (0.0%) | 303 (38.4%) | 498 (26.0%) |
| Not applicable | 722 (100.0%) | 4199 (100.0%) | 0 (0.0%) | 0 (0.0%) |
| **Year group** |  |  |  |  |
| 1 | 0 (0.0%) | 0 (0.0%) | 174 (22.1%) | 444 (23.1%) |
| 2 | 0 (0.0%) | 0 (0.0%) | 131 (16.6%) | 411 (21.4%) |
| 3 | 0 (0.0%) | 0 (0.0%) | 94 (11.9%) | 318 (16.6%) |
| 4+ | 0 (0.0%) | 0 (0.0%) | 390 (49.4%) | 745 (38.8%) |
| Not applicable | 722 (100.0%) | 4199 (100.0%) | 0 (0.0%) | 0 (0.0%) |
| **Household size** |  |  |  |  |
| 1 | 214 (29.6%) | 1177 (28.0%) | 167 (21.2%) | 334 (17.4%) |
| 2-3 | 306 (42.4%) | 1804 (43.0%) | 248 (31.4%) | 647 (33.7%) |
| 4-5 | 140 (19.4%) | 895 (21.3%) | 213 (27.0%) | 589 (30.7%) |
| 6-9 | 4 (0.6%) | 12 (0.3%) | 111 (14.1%) | 259 (13.5%) |
| 10+ | 4 (0.6%) | 7 (0.2%) | 27 (3.4%) | 59 (3.1%) |
| Unknown | 54 (7.5%) | 304 (7.3%) | 23 (2.9%) | 30 (1.6%) |
| **Student residence** |  |  |  |  |
| Catered halls | 0 (0.0%) | 0 (0.0%) | 24 (3.0%) | 60 (3.1%) |
| Self-catered halls | 0 (0.0%) | 0 (0.0%) | 158 (20.0%) | 380 (19.8%) |
| Shared house/flat | 0 (0.0%) | 0 (0.0%) | 412 (52.2%) | 1092 (57.0%) |
| Live with family | 0 (0.0%) | 0 (0.0%) | 81 (10.3%) | 171 (8.9%) |
| Live alone | 0 (0.0%) | 0 (0.0%) | 58 (7.4%) | 117 (6.1%) |
| Other | 0 (0.0%) | 0 (0.0%) | 56 (7.1%) | 97 (5.1%) |
| Not applicable | 722 (100.0%) | 4199 (100.0%) | 0 (0.0%) | 0 (0.0%) |
|  | **N (%) participants first filling out the survey by period** | **N (%) responses for each period** | **N (%) participants first filling out the survey by period** | **N (%) responses for each period** |
| **COVID-19 guidance period** |  |  |  |  |
| 23<sup>rd</sup> June – 3<sup>rd</sup> July 2020 | 329 (45.6%) | 353 (8.4%) | 99 (12.6%) | 68 (3.5%) |
| 4<sup>th</sup> July – 13<sup>th</sup> September 2020 | 359 (49.7%) | 1594 (38.0%) | 64 (8.1%) | 202 (10.5%) |
| 14<sup>th</sup> September – 4<sup>th</sup> November 2020 | 31 (4.3%) | 1623 (38.7%) | 615 (78.0%) | 1314 (68.5%) |
| 5<sup>th</sup> November – 25<sup>th</sup> November 2020 | 3 (0.4%) | 628 (15.0%) | 11 (1.4%) | 333 (17.4%) |

### Variation in contacts over time

Figure 1 shows the mean, median, and IQR of the number of contacts reported on the previous day, stratified by week. For staff, among whom there were high response numbers throughout the entire analysis period, the median number of contacts rose from 2 during the post-first lockdown period to 3 during the relaxed guidance and “rule-of-six” periods and reduced to 2 during the second lockdown period. Similarly, the mean number of daily contacts for staff rose from 3.2 (95% confidence interval [95%CI]: 2.8-3.5) during the post-first lockdown period, to 4.4 (95%CI: 3.9-4.9) during the relaxed guidance period, 5.4 (95%CI: 4.6-6.1) during the “rule-of-six” period and dropped to 3.3 (95%CI: 2.8-3.8) during the second lockdown period.

**Figure 1:**
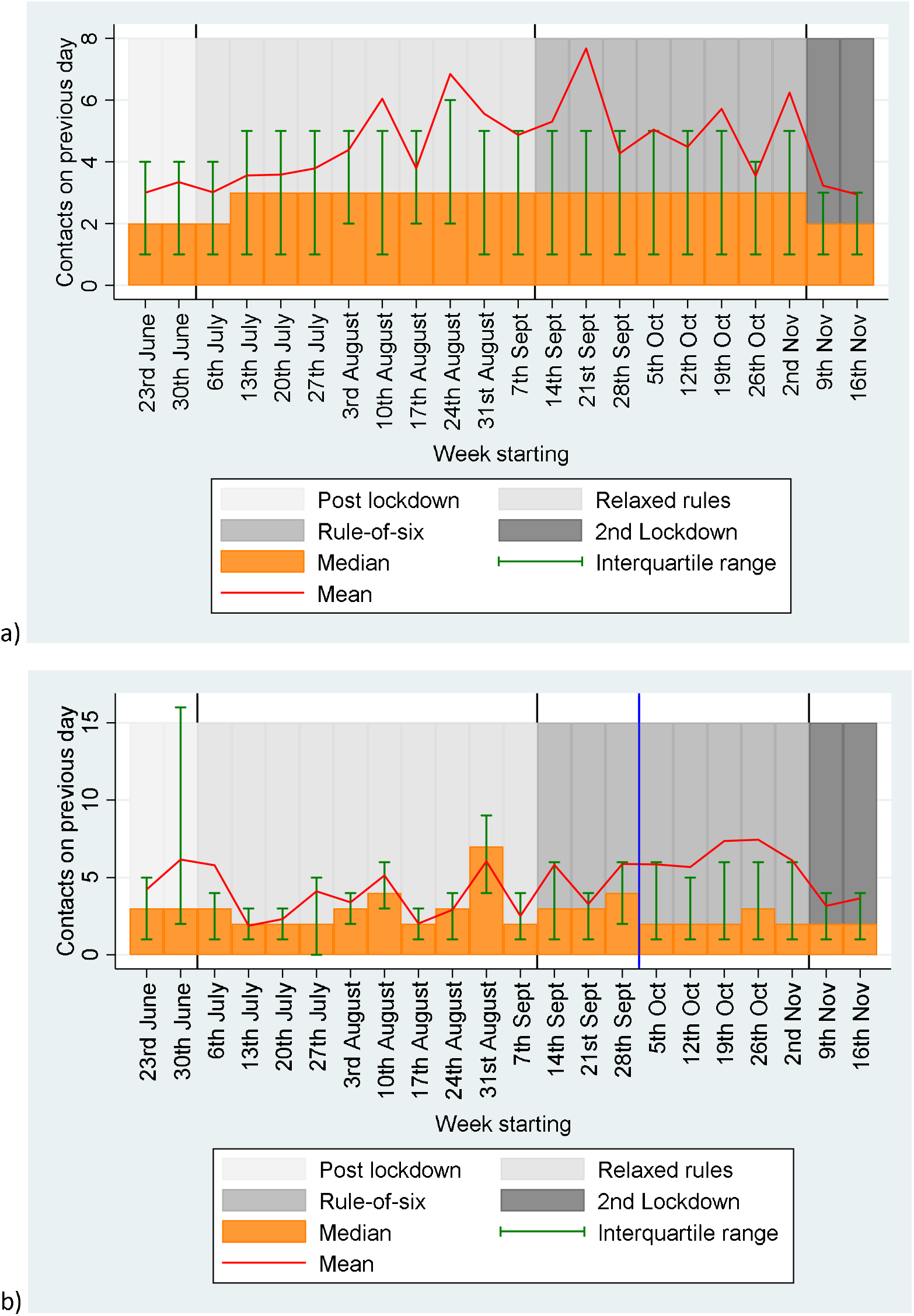
Weighted mean and median (with interquartile ranges) number of contacts for the previous day, stratified by week for a) staff; and b) students. For students the blue line indicates the start of the mass communications campaign*. * Mass communications campaign for students began the week of the 5^th^ October and ended in November

For students, after 05/10/2020, when there were high numbers of responses leading to clearer interpretation, the median daily contacts was 2 and the mean was around 6.2 (95%CI: 5.5-6.9), until the introduction of the second lockdown when it dropped to 4.0 (95%CI: 3.3-4.7).

For both staff and students there was a large difference in the mean and median contacts, as some individuals had large numbers of contacts (see figure 2). Supplementary table 2 shows that there were lower numbers of survey responses at the weekend, but the reported number of contacts was similar for each day. Supplementary figure 1 shows a histogram of contacts, stratified by staff and students.

**Figure 2:**
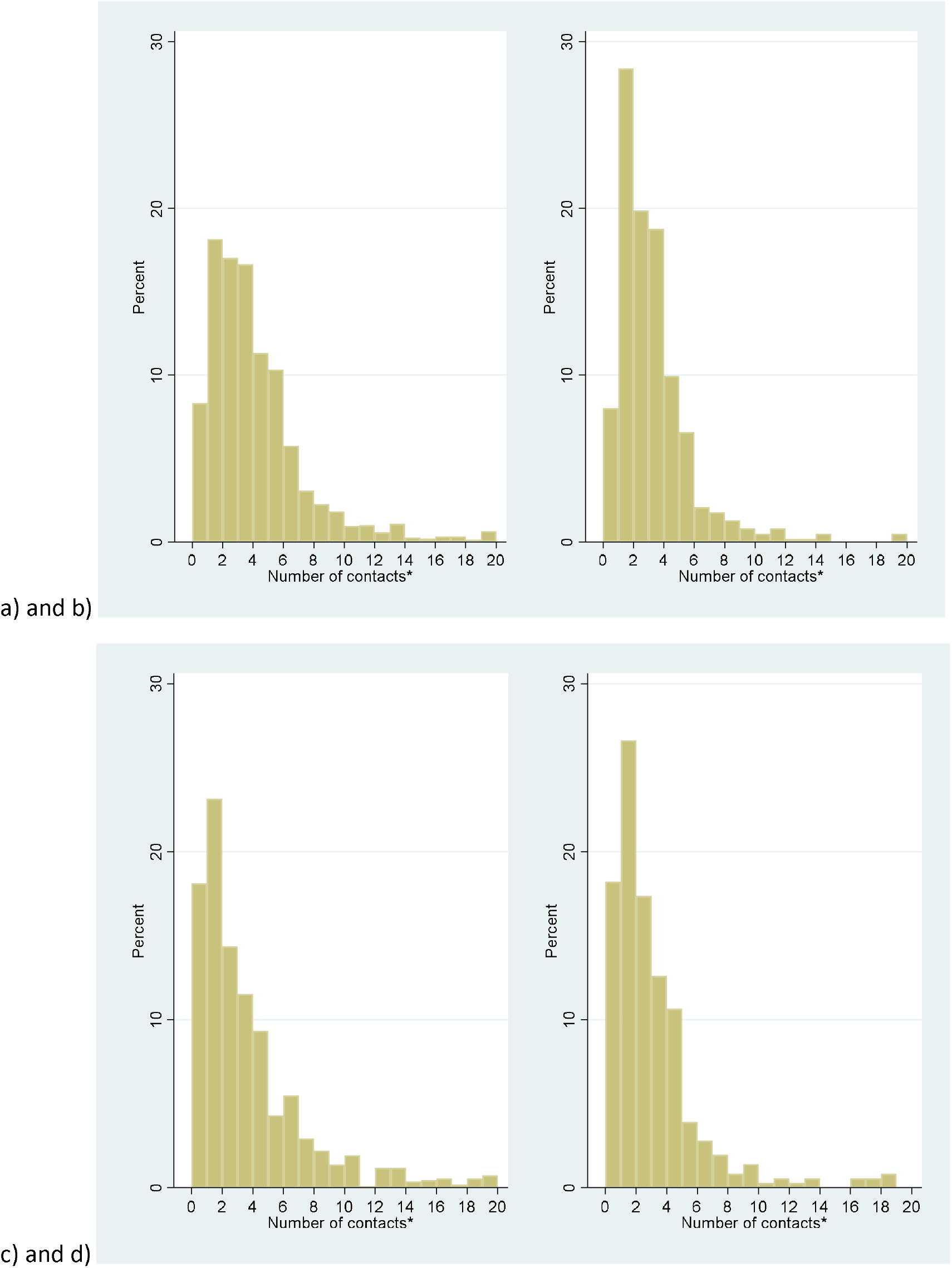
Weighted histograms of the number of contacts on the previous day for staff in a) the “rule-of-six” period (14^th^ Sept-4^th^ Nov) before the second lockdown; and b) for the second lockdown period (5^th^ Nov – 25^th^ Nov); and the same graphs, respectively for students: c) and d). *There were 60/1659 records for staff in the “rule-of-six” period with more than 20 contacts, 11/635 in the second lockdown period, whilst for students there were 78/1171 in the “rule-of-six” period, and 6/363 in the second lockdown period.

### Contacts in “Rule-of-six” period versus second lockdown

Figure 2 shows that there was a shift towards higher proportions of both staff and students having lower contacts in the second lockdown period than in the “rule-of-six” period. Table 3 compares the number of contacts and types of these contacts for staff and students during the “rule-of-six” and second lockdown periods. For staff, the mean overall contacts dropped from 5.4 to 3.3, with a large part of this drop being driven by group contacts falling from a mean of 2.1 to 0.7 (this includes those with 0 group contacts). The mean individual contacts of staff dropped from 2.8 to 2.3, but there was a similar number of these contacts involving touch in both periods (1.4 and 1.3), similar mean numbers of household member contacts (1.4 and 1.4), frequent contacts (1.5 and 1.5), and contacts made at home (1.6 and 1.7). Staff had similar numbers of contacts made at the university over both periods (means 0.5 and 0.5) and similar numbers of UoB contacts (0.8 and 0.7). The mean number of contacts made at locations other than home and university dropped for staff between the two periods, from 2.9 to 1.2.

**Table 3:**
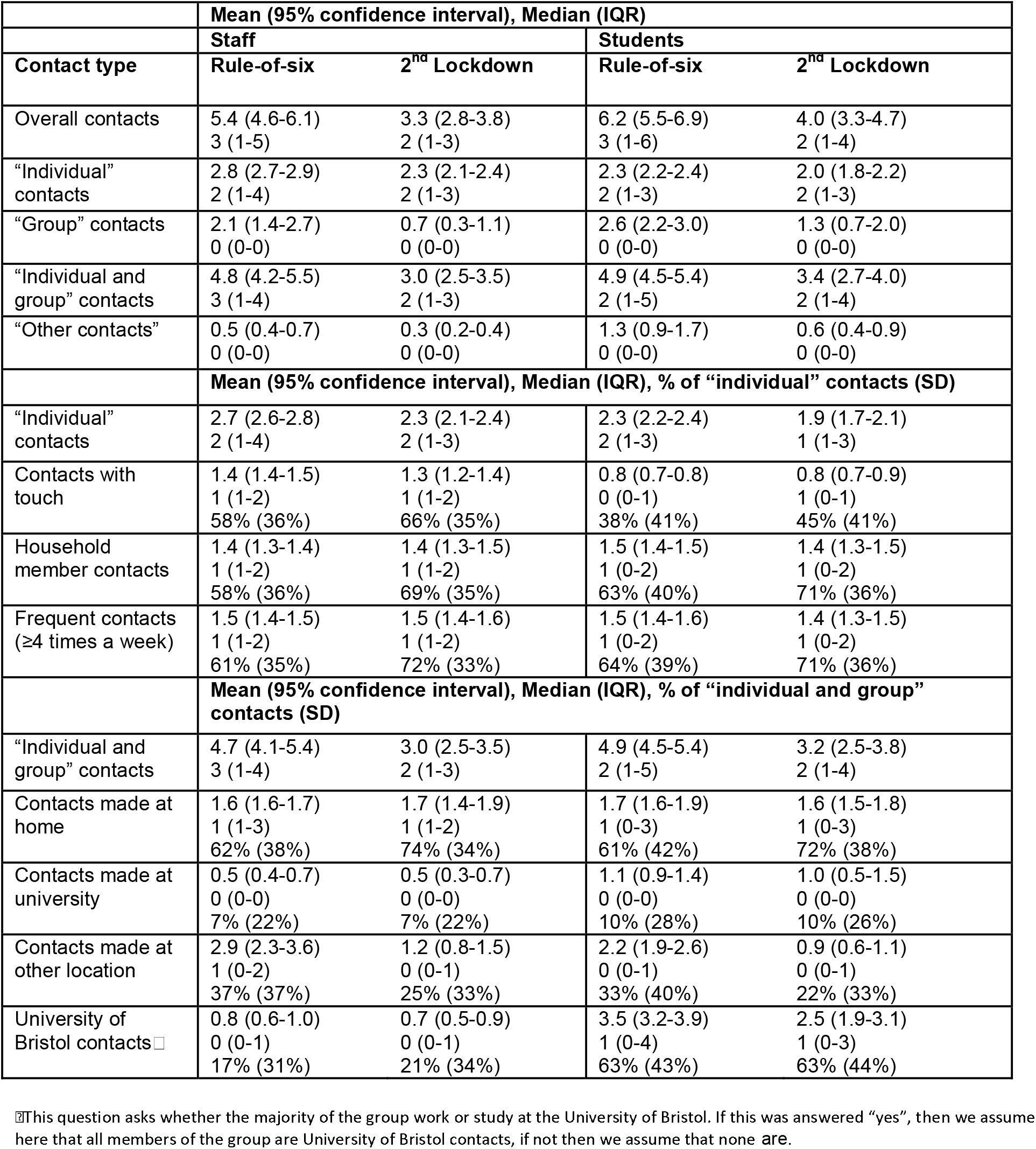
Overall weighted number of contacts on the previous day and types of contacts for “rule-of-six” and second lockdown COVID-19 restriction guidance periods, stratified by staff and students. *“Individual” contacts were the people that the participant spoke to in person one-on-one, including those in the participant’s household and support bubble. “Group” contacts were the contacts that the participant had with large groups of individuals in the same setting (for example, sports teams, tutorials, lectures, religious services, large gatherings with friends and family). “Other” contacts were the many people participants spoke to one-on-one in the same setting where the contacts did not have the opportunity to speak to each other (for example, as part of a customer service role in a shop). Not all of the contact types were asked for each category of contacts, so are only comparable to the associated categories indicated here.

For students, the mean overall number of contacts dropped from 6.3 during the “rule-of-six” period to 4.0 during the second lockdown. Between these two periods, mean individual contacts dropped slightly from 2.3 to 2.0, group contacts dropped from 2.6 to 1.3, and other contacts dropped from 1.3 to 0.6. The mean number of student contacts involving touch was lower than for staff but was consistent across both periods (0.8 and 0.8). Students reported a similar mean number of household member contacts over both periods (1.5 and 1.4) as staff, as well as similar numbers of frequent contacts (1.5 and 1.4), and contacts made at home (1.7 and 1.6). Students had higher mean numbers of contacts made at the university across the two periods than staff (1.1 and 1.0). Students also had higher mean numbers of UoB contacts than staff, however, these dropped between the two periods from 3.5 to 2.5, whilst contacts at locations other than home or university were lower than for staff and dropped between the two periods from 2.2 to 0.9.

### Groups larger than 6

Figure 3 shows the proportion of respondents that met with groups larger than 6 on the previous day for each guidance period. For staff the proportion was lowest in the post-first lockdown period (0.01; 95% confidence interval [95%CI]: 0.00-0.02) and then rose in the relaxed guidance period (0.03; 95%CI: 0.02-0.04) and again in the “rule-of-six” period (0.06; 95%CI: 0.05-0.07), before falling during the second lockdown (0.03; 95%CI: 0.01-0.04). For students, there is large uncertainty in the first two periods due to a lack of responses, but the proportion reporting meeting with groups larger than 6 dropped between the “rule-of-six” period (0.12; 95%CI: 0.10-0.14) and the second lockdown period (0.07; 95%CI: 0.04-0.10), although this was higher than for staff.

**Figure 3:**
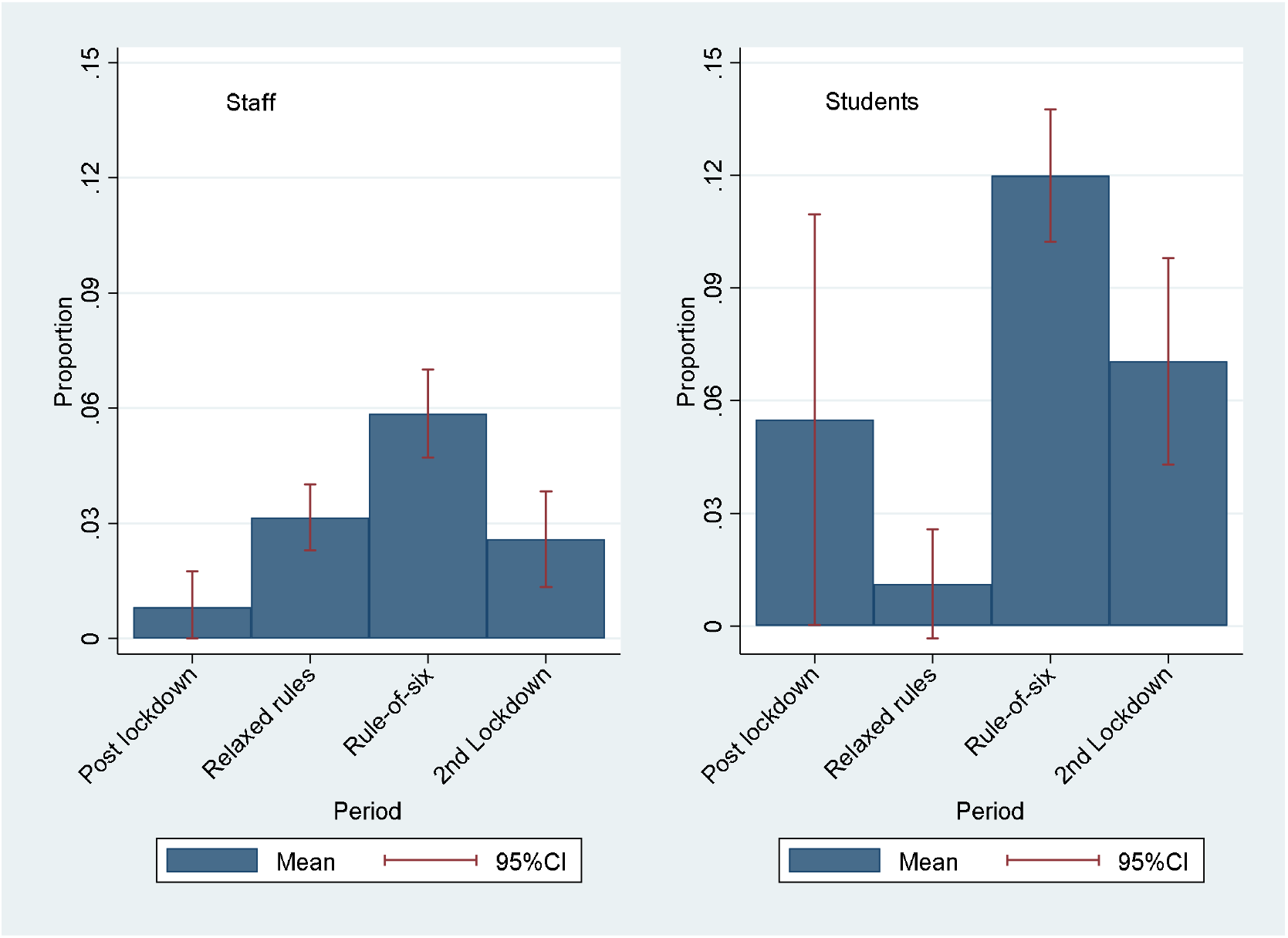
Weighted proportion of respondents that met with groups larger than 6 on the previous day, stratified by staff and students, and by COVID-19 guidance period* *Defined as group contacts of more than 6 for a single group.

### Regression of daily contact numbers

Table 4 contains the multivariable regression analyses results on the number of contacts on the previous day for staff and students. For both staff and students, the number of contacts was higher in the “rule-of-six” period than in the other periods and was lower in weeks they had been isolating.

**Table 4:** Weighted univariable and multivariable regression of the number of contacts on the previous day for a) staff and b) students

| VARIABLE | N: MEAN (95%CI)<br>CONTACTS | UNIVARIABLE |  | MULTIVARIABLE |  |
| --- | --- | --- | --- | --- | --- |
|  |  | IRR (95%CI) | p-value | IRR (95%CI) | p-value |
| POST-FIRST LOCKDOWN PERIOD | 353: 3.2 (2.8-3.5) | 0.59 (0.52-0.67) | <0.001 | 0.62 (0.54, 0.71) | <0.001 |
| RELAXED GUIDANCE PERIOD | 1594: 4.4 (3.9-4.9) | 0.82 (0.76-0.88) | <0.001 | 0.83 (0.77, 0.89) | <0.001 |
| RULE-OF-SIX PERIOD | 1624: 5.4 (4.6-6.1) | Reference | NA | Reference | NA |
| 2<sup>ND</sup> LOCKDOWN PERIOD | 628: 3.3 (2.8-3.8) | 0.62 (0.55-0.68) | <0.001 | 0.64 (0.58, 0.71) | <0.001 |
| AGE 17-24 | 31: 4.8 (2.7-6.9) | 1.06 (0.72-1.56) | 0.773 | 1.02 (0.69, 1.52) | 0.910 |
| AGE 25-44 | 2116: 4.5 (3.9-5.1) | Reference | NA | Reference | NA |
| AGE 45-64 | 1915: 4.6 (4.2-5.0) | 1.02 (0.95-1.09) | 0.619 | 1.00 (0.93, 1.07) | 0.932 |
| AGE 65-79 | 112: 2.8 (2.4-3.2) | 0.63 (0.50-0.78) | <0.001 | 0.76 (0.60, 0.96) | 0.020 |
| AGE ≥80 | 25: 1.8 (1.3-2.3) | 0.39 (0.24-0.64) | <0.001 | 0.46 (0.27, 0.78) | 0.004 |
| FEMALE/OTHER | 2309: 4.7 (4.2-5.2) | Reference | NA | Reference | NA |
| MALE | 1990: 4.2 (3.7-4.7) | 0.89 (0.83-0.95) | 0.001 | 0.91 (0.85, 0.98) | 0.009 |
| NO SYMPTOMS LAST WEEK | 3252: 4.3 (3.9-4.6) | Reference | NA | Reference | NA |
| SYMPTOMS LAST WEEK | 946: 5.3 (4.3-6.4) | 1.26 (1.16-1.36) | <0.001 | 1.20 (1.10, 1.30) | <0.001 |
| NO CARDINAL SYMPTOMS LAST WEEK | 4139: 4.5 (4.2-4.9) | Reference | NA | Reference | NA |
| CARDINAL SYMPTOMS LAST WEEK | 60: 3.0 (1.9-4.2) | 0.67 (0.50-0.90) | 0.008 | 0.65 (0.47, 0.90) | 0.009 |
| NOT ISOLATED LAST WEEK | 4103: 4.5 (4.2-4.9) | Reference | NA | Reference | NA |
| ISOLATED LAST WEEK | 96: 2.3 (1.9-2.7) | 0.50 (0.40-0.64) | <0.001 | 0.49 (0.38, 0.63) | <0.001 |
| NOT HIGH RISK | 3935: 4.5 (4.1-4.8) | Reference | NA | Reference | NA |
| HIGH RISK | 264: 5.1 (2.4-7.8) | 1.14 (1.00-1.31) | 0.056 | 1.29 (1.11, 1.50) | 0.001 |
| HOUSEHOLD SIZE 1 | 1177: 3.1 (2.7-3.5) | 0.67 (0.61-0.73) | <0.001 | 0.68 (0.62, 0.74) | <0.001 |
| HOUSEHOLD SIZE 2-3 | 1804: 4.6 (4.2-5.1) | Reference | NA | Reference | NA |
| HOUSEHOLD SIZE 4-5 | 894: 6.3 (5.4-7.1) | 1.35 (1.24-1.47) | <0.001 | 1.35 (1.24, 1.48) | <0.001 |
| HOUSEHOLD SIZE 6-9 | 12: 2.9 (1.9-3.9) | 0.63 (0.33-1.22) | 0.173 | 0.75 (0.39, 1.46) | 0.403 |
| HOUSEHOLD SIZE 10+ | 7: 1.4 (0.0-3.1) | 0.30 (0.11-0.82) | 0.019 | 0.36 (0.13, 0.99) | 0.047 |
| HOUSEHOLD SIZE MISSING | 304: 4.0 (1.4-6.5) | 0.85 (0.74-0.98) | 0.022 | 0.78 (0.68, 0.91) | 0.001 |
| NO COVID-19 | 3621: 4.5 (4.1-4.8) | Reference | NA | Reference | NA |
| PREVIOUSLY TESTED POSITIVE MORE THAN 2 WEEKS BEFORE SURVEY | 22: 1.2 (1.1-1.4) | 0.28 (0.16-0.49) | <0.001 | 0.43 (0.24, 0.77) | 0.005 |
| PREVIOUSLY SUSPECTED TO BE POSITIVE MORE THAN 2 WEEKS | 507: 4.9 (3.4-6.4) | 1.10 (0.99-1.22) | 0.063 | 1.08 (0.97, 1.20) | 0.169 |
| SUSPECTED TO BE POSITIVE IN LAST 2 WEEKS | 46: 3.8 (2.0-5.6) | 0.84 (0.61-1.17) | 0.312 | 0.95 (0.67, 1.34) | 0.766 |
| TESTED POSITIVE IN LAST 2 WEEKS | 4: 2.1 (1.1-3.1) | 0.47 (0.14-1.54) | 0.214 | 1.03 (0.30, 3.54) | 0.957 |

b)
| VARIABLE | N: MEAN (SD)<br>CONTACTS | UNIVARIABLE |  | MULTIVARIABLE |  |
| --- | --- | --- | --- | --- | --- |
|  |  | IRR (95%CI) | p-value | IRR (95%CI) | p-value |
| POST-FIRST LOCKDOWN PERIOD | 68: 4.5 (3.2-5.8) | 0.72 (0.55-0.95) | 0.018 | 0.87 (0.65-1.15) | 0.316 |
| RELAXED GUIDANCE PERIOD | 202: 3.8 (2.6-5.0) | 0.61 (0.52-0.72) | <0.001 | 0.70 (0.59-0.84) | <0.001 |
| RULE-OF-SIX PERIOD | 1314: 6.2 (5.5-6.9) | Reference | NA | Reference | NA |
| 2<sup>ND</sup> LOCKDOWN PERIOD | 333: 4.0 (3.3-4.7) | 0.64 (0.56-0.73) | <0.001 | 0.60 (0.52-0.69) | <0.001 |
| AGE 17-24 | 1499: 5.9 (5.3-6.6) | Reference | NA | Reference | NA |
| AGE 25-44 | 386: 4.1 (3.2-5.0) | 0.69 (0.61-0.78) | <0.001 | 0.89 (0.73-1.10) | 0.280 |
| AGE 45-64 | 24: 2.9 (1.5-4.3) | 0.49 (0.31-0.78) | 0.003 | 0.50 (0.29-0.84) | 0.010 |
| AGE 65-79 | 7: 2.6 (0.0-5.6) | 0.44 (0.18-1.06) | 0.068 | 0.75 (0.28-1.98) | 0.559 |
| AGE ≥80 | 2: 1.0 (1.0-1.0) | 0.17 (0.01-3.10) | 0.231 | 0.32 (0.02-6.09) | 0.171 |
| FEMALE/OTHER | 1064: 4.8 (4.3-5.3) | Reference | NA | Reference | NA |
| MALE | 853: 6.4 (5.5-7.4) | 1.33 (1.21-1.47) | <0.001 | 1.14 (1.02-1.27) | 0.017 |
| UNDERGRAD | 1419: 6.1 (5.5-6.7) | Reference | NA | Reference | NA |
| POSTGRAD | 498: 4.0 (3.1-4.9) | 0.66 (0.59-0.74) | <0.001 | 0.56 (0.45-0.71) | <0.001 |
| STUDY YEAR 1 | 444: 4.2 (3.5-4.9) | Reference | NA | Reference | NA |
| STUDY YEAR 2 | 411: 7.2 (6.1-8.2) | 1.70 (1.47-1.97) | <0.001 | 1.62 (1.62-2.49) | <0.001 |
| STUDY YEAR 3 | 318: 4.6 (3.7-5.5) | 1.10 (0.93-1.28) | 0.262 | 1.30 (1.03-1.65) | 0.027 |
| STUDY YEAR 4+ | 745: 5.8 (4.8-6.8) | 1.38 (1.21-1.57) | <0.001 | 2.23 (1.80-2.76) | <0.001 |
| NO SYMPTOMS LAST WEEK | 1301: 5.0 (4.5-5.5) | Reference | NA | Reference | NA |
| SYMPTOMS LAST WEEK | 616: 6.6 (5.5-7.7) | 1.32 (1.18-1.46) | <0.001 | 1.23 (1.09-1.38) | 0.001 |
| NO CARDINAL SYMPTOMS LAST WEEK | 1796: 5.3 (4.9-5.8) | Reference | NA | Reference | NA |
| CARDINAL SYMPTOMS LAST WEEK | 121: 8.3 (4.1-12.5) | 1.55 (1.28-1.89) | <0.001 | 1.24 (0.97-1.60) | 0.090 |
| NOT ISOLATED LAST WEEK | 1690: 5.7 (5.2-6.3) | Reference | NA | Reference | NA |
| ISOLATED LAST WEEK | 227: 4.2 (3.0-5.4) | 0.73 (0.63-0.85) | <0.001 | 0.62 (0.51-0.74) | <0.001 |
| NOT HIGH RISK | 1740: 5.6 (5.1-6.1) | Reference | NA | Reference | NA |
| HIGH RISK | 176: 4.9 (3.2-6.6) | 0.87 (0.73-1.03) | 0.110 | 1.00 (0.83-1.20) | 0.973 |
| HOUSEHOLD SIZE 1 | 334: 4.2 (2.9-5.6) | 0.70 (0.60-0.81) | <0.001 | 1.15 (0.96-1.38) | 0.138 |
| HOUSEHOLD SIZE 2-3 | 647: 6.0 (5.0-7.1) | Reference | NA | Reference | NA |
| HOUSEHOLD SIZE 4-5 | 589: 6.0 (5.2-6.7) | 0.98 (0.88-1.11) | 0.790 | 1.13 (0.98-1.29) | 0.088 |
| HOUSEHOLD SIZE 6-9 | 259: 5.6 (4.5-6.7) | 0.93 (0.80-1.08) | 0.339 | 1.03 (0.86-1.24) | 0.722 |
| HOUSEHOLD SIZE 10+ | 59: 3.7 (2.22-5.1) | 0.61 (0.45-0.82) | 0.001 | 1.00 (0.71-1.96) | 0.939 |
| HOUSEHOLD SIZE MISSING | 30: 3.3 (1.1-5.5) | 0.55 (0.36-0.84) | 0.005 | 0.80 (0.52-1.25) | 0.334 |
| NO COVID-19 | 1508: 5.2 (4.6-5.8) | Reference | NA | Reference | NA |
| PREVIOUSLY TESTED POSITIVE MORE THAN 2 WEEKS BEFORE SURVEY | 34: 7.0 (4.1-9.8) | 1.34 (0.93-1.92) | 0.118 | 1.26 (0.87-1.84) | 0.223 |
| PREVIOUSLY SUSPECTED TO BE POSITIVE MORE THAN 2 WEEKS | 249: 7.8 (6.1-9.6) | 1.51 (1.30-1.74) | <0.001 | 1.38 (1.18-1.60) | <0.001 |
| SUSPECTED TO BE POSITIVE IN LAST 2 WEEKS | 75: 4.8 (3.3-6.3) | 0.92 (0.72-1.19) | 0.549 | 1.08 (0.82-1.42) | 0.580 |
| TESTED POSITIVE IN LAST 2 WEEKS | 56: 3.4 (2.4-4.4) | 0.65 (0.47-0.89) | 0.007 | 0.78 (0.52-1.17) | 0.226 |
| CATERED HALLS | 60: 2.3 (1.5-3.0) | 0.38 (0.28-0.52) | <0.001 | 0.65 (0.45-0.95) | 0.025 |
| SELF-CATERED HALLS | 380: 6.2 (4.6-7.8) | 1.04 (0.92-1.18) | 0.519 | 1.25 (1.02-1.54) | 0.032 |
| SHARED HOUSE/FLAT | 1092: 5.9 (5.3-6.5) | Reference | NA | Reference | NA |
| LIVE WITH FAMILY | 171: 5.7 (3.7-7.7) | 0.96 (0.81-1.15) | 0.687 | 1.36 (1.10-1.67) | 0.005 |
| **LIVE ALONE** | 117: 1.9 (1.2-2.6) | 0.2 (0.25-0.40) | <0.001 | 0.42 (0.31-0.56) | <0.001 |
| **OTHER** | 97: 4.3 (2.8-5.8) | 0.72 (0.57-0.90) | 0.004 | 1.07 (0.82-1.39) | 0.631 |

IRR: Incidence rate ratio. SD: Standard deviation. CI: Confidence interval.

Table 4a contains the results of the regression analyses on the number of contacts on the previous day for staff. In multivariable analysis, the number of contacts was higher in the “rule-of-six” period than in the other periods. Being aged over 65 was associated with a lower number of contacts in comparison with the 25-44 age group, whilst males reported lower numbers of contacts than females (adjusted incidence rate ratio [aIRR] 0.91; 95% confidence interval [95%CI]: 0.85-0.98). Staff reporting symptoms during the previous week had a higher number of contacts on the previous day than those without symptoms, aIRR 1.20 (95%CI: 1.10-1.30), whilst those with cardinal symptoms had fewer contacts, aIRR 0.65 (95%CI: 0.47-0.90). Staff that had been isolating during the previous week had lower contacts on the previous day, aIRR 0.49 (95%CI: 0.38-0.63), whilst staff that were in high-risk health status groups had higher contacts, aIRR 1.29 (95%CI: 1.11-1.50). Compared with staff living in a household of 2-3 people, staff with a household size of 1 had fewer contacts, aIRR 0.68 (95%CI: 0.62-0.74), whilst staff with a household of 4-5 people had more contacts, aIRR 1.35 (95%CI: 1.24-1.48). Compared with staff that had never tested positive or thought they had never been positive, staff that had previously tested positive more than 2 weeks before the survey had lower numbers of contacts on the previous day, aIRR 0.43 (95%CI: 0.24-0.77).

For students, the regression analysis results are shown in Table 4b. Similarly to staff, the “rule-of-six” period was associated with a higher number of contacts on the previous day than the other periods in the multivariable analysis. Higher age was associated with a lower number of contacts on the previous day. Unlike for staff, males had a higher number of contacts on the previous day than females, aIRR 1.14 (95%CI: 1.02-1.27). Postgraduates reported a lower number of contacts than undergrads, aIRR 0.56 (95%CI: 0.45-0.71), whilst students in study year 1 had lower numbers of contacts than students in other years. As with staff, students reporting symptoms during the last week had higher numbers of contacts on the previous day than those not reporting symptoms, aIRR 1.23 (95%CI: 1.09-1.38), and those isolating during the last week had fewer contacts than those that had not been isolating, aIRR 0.62 (95%CI: 0.51-0.74). For students, there was no difference in daily contacts by household size. Students living in catered halls reported fewer contacts on the previous day than those living in a shared house/flat, aIRR 0.65 (95%CI: 0.45-0.95), whilst those living with their family had higher contacts than those in shared houses/flats, aIRR 1.36 (95%CI: 1.10-1.67). Students living alone had lower numbers of contacts than those living in a shared house/flat, aIRR 0.42 (0.31-0.56). Students that previously suspected themselves to be positive more than 2 weeks before taking the survey reported higher numbers of contacts on the previous day than those that had never tested positive nor suspected themselves of having COVID-19, aIRR 1.38 (95%CI: 1.18-1.60).

## Discussion

For both the university staff and students, the number of contacts on the previous day was higher in the “rule-of-six” period than in the post-first lockdown period, the relaxed guidance period, and the second lockdown.

For staff, contacts remained low throughout the analysis period, rising between the post-first lockdown period (median: 2, mean: 3.2), the relaxed guidance period (median: 3, mean: 4.4), the “rule-of-six” period (median: 3, mean: 5.4), and dropping during the second lockdown (median: 2, mean: 3.3). The difference between the median and means due to some individuals reporting many contacts. The drop in mean contacts between the last two periods for staff was mostly driven by a mean reduction in contacts in locations other than home or university (from 2.9 to 1.2), including group contacts (from 2.1 to 0.7), whilst there was a similar number of household member contacts between both periods (1.4 and 1.4) and those made at the university (0.5 and 0.5). This indicates that staff members reduced their numbers of social contacts and mostly remained in contact with their household members.

For students, there were few responses until October when a mass communications campaign was launched, after which, the number of contacts on the previous day remained low, the median was 2 and the mean was 6.2 during the “rule-of-six” period, dropping to 4.0 in the second lockdown.

The lower median contacts during the early weeks of term for students than staff was perhaps due to a high percentage of students having to isolate: both students and staff that were isolating had lower numbers of contacts than those not isolating. The drop in mean number of contacts for students between the last two periods was driven by a reduction in all contact types except for those made at home (1.7 to 1.6), which, similarly to staff, indicated a reduction in social contacts. Students also had higher mean numbers of UoB contacts than staff, however, for students these dropped between the two last periods from 3.5 to 2.5.

For both staff and students, the proportion meeting with groups larger than 6 dropped between the “rule-of-six” period and the second lockdown period, although was higher for students than for staff. A study^14^ suggests that in the COVID-19 pandemic, in contrast to previous research on adherence to non-pharmaceutical interventions in a pandemic^15^, that there have been high levels of adherence even when individuals believe themselves to be at comparatively low risk from the disease to other groups. This is seen in our study where students were highly compliant with the regulations during various COVID-19 regulation periods, despite most students being in a low-risk age group. Where students were meeting with groups larger than six during the rule of six period; this could have been due to exemptions for sports groups, teaching group sizes or living situations. Alternatively, these could reflect non-adherence to regulations, with the main barriers to adherence in students having been previously identified as a fear of mental health impacts and loneliness^16^. It must be noted that compliance related to hygiene has been found to be uniformly distinct from compliance related to social distancing behaviours and that treating public health compliance as one construct can lead to poorer prediction of compliance behaviour and poorer production of effective recommendations for public health^14^. Therefore, the compliance to social distancing regulations we found here may not indicate that there has been similar compliance to hygiene practices in staff and students during the COVID-19 pandemic.

### Comparison with other literature

For each guidance period studied we found a lower mean number of daily contacts among our staff and student populations than was found in the pre-COVID-19 era Warwick social contacts survey from 2009^17,18^, either among their entire sample (26.8) or the students in that sample (29.9). The students in the Warwick survey had more home contacts (3.5) than other participants (2.3), whilst most contacts for students (82%, 95%CI: 79%-86%) were either at home or university-related. Students reported 20 (95%CI: 14.1-28.8) university-related contacts. Similarly, we found that a high percentage of the contacts of students were either at home or university (∼72%) and that our staff (comparing with the Warwick survey’s “other participants”) had 1.6 home contacts. However, we found that students had a daily mean of 1.7 contacts at home and 2.5 university contacts, possibly indicating that the national and university guidance was successful in reducing contacts. Meanwhile, the POLYMOD social contacts survey^19^ found a lower mean than Warwick social contacts survey (11.7) in their Great Britain sample (average age ∼30), but still much higher than the mean values we recorded for either staff or students. The BBC Pandemic project reported number of daily contacts from a national study in 2018, with a mean of 10.5^20^, also much higher than we reported.

The CoMix study found during the first COVID-19 lockdown 24-29 March that mean contacts were 2.8 among their general population participants^21^, comparable to the 3.0 contacts among staff during the 2^nd^ lockdown period in our study. The COVID-19 Contact Network (CoCoNet) Study was conducted between 28 July and 14 August in the general population, with preliminary findings suggesting a mean of 2.9 daily non-household contacts per person^22^. Similarly, we report 0.5 contacts in university among staff and 2.9 in non-home, non-university settings in the “rule-of-six” period.

### Strengths and limitations

The strengths of this survey include the sample size, longitudinal format, and anonymous nature that enabled us to capture self-reported contact patterns of a large number of staff and students during a key period in the UK’s COVID-19 pandemic. It provides a unique data source on student and staff behaviour during the pandemic for informing public health action and mathematical models. Results for students are likely generalisable to other UK city-based universities, and to some city-based universities in other countries. Meanwhile, the staff results are likely generalisable to a working cohort of the general population, due to their age profile. Survey questions were designed to be comparable to existing contact surveys^17-19^.

However, the survey started after the first lockdown period, so we are unable to compare whether contacts during the second lockdown were higher than in the first. Also, we cannot ascertain what caused the changes in numbers of contacts. We lack student data for the early period of the survey, as data collection could not be scaled up until October, therefore, we only have robust data on students from October onwards. Additionally, those with many contacts or with little available time may have been deterred from completing it, which may mean it is not representative. We include clear instructions defining “contacts” in the survey; however, people may interpret the instructions differently leading to variation in what people considered a contact to be.

Selection bias for people particularly engaged in health-seeking behaviours may have occurred. However, we did capture individuals reporting large numbers of contacts. There will inevitably be issues regarding recall bias, and issues with response bias, leading to inaccurate or false responses.

### Implications

This study comes at a unique time when a lockdown has been implemented to reduce contacts between individuals. However, the number of reported daily cases of COVID-19 is still high^6^. Bristol went into the second lockdown covered by this study in the lowest tier of COVID-19 restrictions and came out (as with much of the country) in the highest tier, with a third lockdown then implemented in 2021^4^. UoB, as with many other UK universities, is preparing to manage a possible mass migration events of its students back to university when the current lockdown is relaxed, with the potential for COVID-19 transmission to escalate due to enhanced population mixing^23^. The setting is important due to its uniqueness, as universities were allowed to carry on teaching throughout the lockdown, meaning that some mixing between households still occurred^3^, whilst the setting is also generalisable, as university staff are likely relatively representative of many working-age populations in age structure, enabling us to estimate the difference in contacts between students and the general population. It is important to be able to understand the effect of the COVID-19 guidance changes, particularly lockdowns, on people’s behaviour for any future pandemics that could occur. We show that on average there was high adherence to the guidance throughout the survey period for both staff and students, despite students receiving negative media coverage during the pandemic^24^. The average number of contacts remained low throughout the study and few people were meeting groups larger than 6, despite many students living in large households and attending lectures. Students had slightly higher numbers of overall contacts than staff during, however, there was a reduction in the number of contacts during the second lockdown for both groups, returning them to the levels in the period after the first UK lockdown, primarily driven by a reduction in social contacts, whilst daily household contact numbers remained steady.

## Data Availability

Data are available from the corresponding author.

## Acknowledgements

We would like to thank the Elizabeth Blackwell Institute for funding this research, our RedCap data manager Alison Horne and our PPI group for their feedback during the development of the survey. We would also like to thank all the participants who have taken part in this study.

