## Supplementary materials for "University students and staff able to maintain low daily contact numbers during various COVID-19 guideline periods"

*Further survey information*

Participants who had signed up to repeat questionnaires were sent an email every 14 days (changing to every 8 days from 13/09/2020 onwards to capture more information ahead of the return to campus of the students) with a unique link that allowed their responses to be anonymously connected to those from previous CON-QUEST questionnaires that they had responded to. The reminder emails with the survey links were sent regardless of whether participants have filled in surveys from previous reminder emails or when they responded to them.

For “individual” contacts (contact type 1), participants were asked about where this contact was made, whether this contact was indoors, outdoors or both, the duration of this contact, whether this contact involved touch, whether this contact studied/worked at the university (and if so which faculty and school they were associated with), their age, whether they were part of their household, and how often they would usually have contact with this person.

For “other” contacts (contact type 2), no additional questions were asked, as it was expected that there often would be a large number of “other” contacts and participants would not be motivated to answer additional questions on them.

For “group” contacts (contact type 3), participants were asked how many individuals this involved, their ages, whether the majority were from UoB (and if so the main faculty and school this group was associated with), where the group met, whether this was indoors, outdoors or both, whether the members of the group talked to each other and how long the contact with this group was for.

Additionally, participants were asked about symptoms in the last 7 days, whether they had sought medical attention for these symptoms, whether they had been self-isolating in the last 7 days, and their COVID-19 status. For some analyses, the variable on whether people have had COVID-19 (no, yes confirmed by a test, yes a doctor suspected so, yes my own suspicions) was combined with the date that they had been tested or were suspected to have COVID-19. This was to create new variables on whether they had COVID-19 in the two weeks prior to survey completion, or before this.

*Survey amendments on 13/09/2020*

- Rewritten and shortened the introduction with the purpose of making it more accessible and easier to read.
- Updated question on those at higher risk of severe illness to be in line with the names used for at risk groups now used by the NHS
- Added in a further question on job type for staff and expanded the staff faculty and school question to include directorates and institutions. This is to be more inclusive and to distinguish those who could be at higher risk of COVID-19 due to their job type e.g. Staff who work in Operational services can work across multiple buildings and have more contacts with others
- New question on whether working from home is feasible
- Adapted the questions on where students live to make it more relevant and up to date
- Moved the question on which country you are living in and optional postcode question to the repeat questionnaire, since when people complete the repeat questionnaire they might be in different locations and we do not want the new data they are entering on contacts and symptoms to be related to an incorrect location.
- Changed the wording on halls of residence, since there are now going to be official “living circles” which are like households within halls
- Removed question “Is your household made up of different people as a result of the COVID-19 pandemic?” and the follow up question, as well as the questions on “Due to COVID-19 have you, or would you consider taking the following action when the University is open?” since this data is no longer important to collect.
- New question on support bubbles – these were not in practice at the time of releasing the survey, however, this is important contact data to collect.
- Deleted question on willingness to use symptom tracker apps or contact tracing apps as this is being collected in other surveys
- Added in finger prick option for testing question
- Added in question on transport used yesterday
- Clarified time asked about for “Do you think you have had COIVD-19?”
- Clarified time period for question on contact with COVID-19 case in last two weeks
- Some clarifications in the symptoms question
- Clarifications in the instructions for the contacts questions
- Clarification in instructions after survey is submitted
- Some additions of extra words to clarify questions throughout the survey

*Channels for mass communication about the CON-QUEST survey to students*

1^st^ October- Halls inductions email

4^th^ October 2020- Find your Bristol -Twitter

8^th^ October- Find Your Bristol- Twitter and Facebook

14^th^ October- October Student Newsletter

Other communications ongoing in October:

- Find your Bristol- post to portal, email to all students on safety
- Elizabeth Blackwell Institute social media
- Student Union Comms Champions
- Student administration shared in regular newsletters
- Shared with Faculty Marketing group
- Link added to the end of the Students Union welcome survey

**Weighting**

For students, initial analyses suggested males and undergraduates were underrepresented in the survey responses compared to the overall UoB student population. We applied weights during analyses on students, with weights based on publicly available UoB data on student demographics, to make the dataset more representative of the university’s student population – supplementary table 1.

We had separate data available for staff ages and staff gender breakdown so had to choose one. Initial investigations revealed the age breakdown in CONQUEST was more similar to that in the university than the gender breakdown (23.8% male in the survey; 45.0% male in the university data) so weighting by gender was used.

**Supplementary table 1:** Comparison of university data on student demographics with the corresponding CONQUEST survey data demographic information

| **Population** | **University data http://www.bristol.ac.uk/ssio/statistics/** | **CONQUEST data (including multiple records per respondent)** |
| --- | --- | --- |
| Female, undergraduate | 39.6% | 35.0% |
| Female, postgraduate | 15.7% | 34.6% |
| Male, undergraduate | 34.3% | 13.2% |
| Male, postgraduate | 10.5% | 16.6% |

**Number of contacts by day of the week**

**Supplementary table 2:** Number of contacts by day of the week for staff and students (unweighted).

|  | **Mean number of contacts (standard deviation) - N** | |
| --- | --- | --- |
| **Weekday** | **Staff** | **Students** |
| Monday | 5.1 (15.8) – 715 | 5.8 (14.3) – 279 |
| Tuesday | 4.0 (10.0) – 693 | 4.8 (11.4) – 273 |
| Wednesday | 5.0 (12.6) – 654 | 5.5 (10.4) – 306 |
| Thursday | 4.1 (8.5) – 562 | 4.5 (7.4) – 338 |
| Friday | 4.2 (5.7) – 234 | 4.6 (7.3) – 153 |
| Saturday | 5.1 (11.0) – 269 | 4.6 (11.5) – 154 |
| Sunday | 4.6 (11.8) - 1072 | 4.2 (8.1) - 403 |

**Supplementary figure 1: Histogram of the number of contacts on the previous day for a) staff, and b) students.**

**a)**

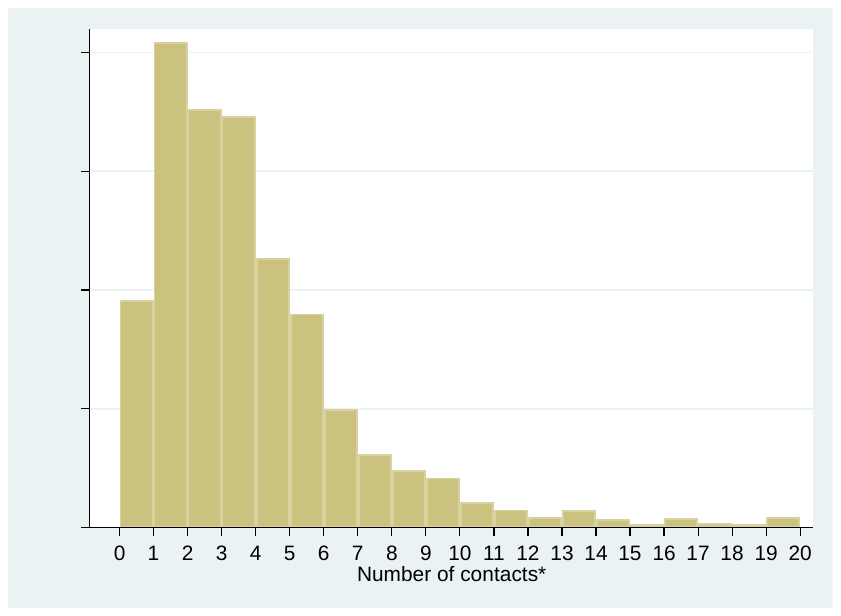

***101 staff members had 21 or more contacts on the previous day.**

**b)**

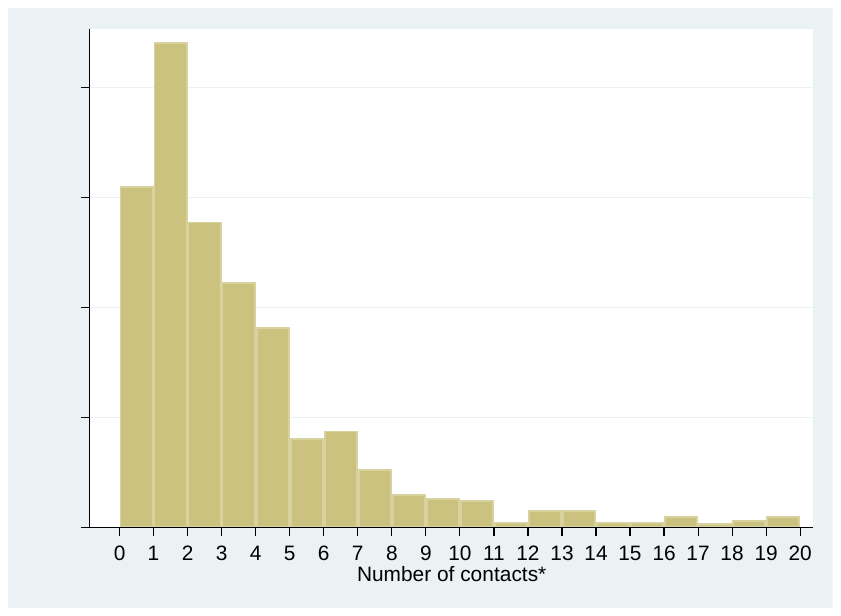

***90 students had 21 or more contacts on the previous day.**

***Description of Tiers in England***

Following the easing of regulations after the first national lockdown, there were various local regulations introduced between July and September 2020, which were replaced on the 14^th^ October 2020 by the tier system. This allocated regions in England into one of three tiers: Tier 1 (medium), Tier 2 (high) and Tier 3 (very high). The regulations in each tier are outlined in full the in the Statutory Instruments (<https://www.legislation.gov.uk/uksi/2020/1103/made>, <https://www.legislation.gov.uk/uksi/2020/1104/made>, <https://www.legislation.gov.uk/uksi/2020/1105/made>) and are summarized here in Supplementary table 3. Bristol remained in Tier 1 from the 14^th^ October until the national lockdown on the 5^th^ November. However, Bristol City Council implemented extra measures implemented in Bristol from 28^th^ October including the appointment of eight “COVID-19 marshalls” to patrol the city’s streets and the council taking on some local responsibility for test and trace. Bristol City Council called this “Tier 1 plus”, however, this name was not officially recognised by the government (Reference: <https://www.bbc.co.uk/news/uk-england-bristol-54721829>)

**Supplementary table 3:** Restrictions for gatherings in different Tiers from 14^th^ October 2020 to 5^th^ November 2020 in England. The “Rule of 6” indicates that no more than 6 people were permitted to participate in a social gathering.

|  | **Tier 1** | **Tier 2** | **Tier 3** |
| --- | --- | --- | --- |
| **Private dwelling (indoors)** | Rule of 6 | Not permitted | Not permitted |
| **Private dwelling (outdoors)** | Rule of 6 | Rule of 6 | Not permitted |
| **Other private indoor space** | Rule of 6 | Not permitted | Not permitted |
| **Other private outdoor space** | Rule of 6 | Rule of 6 | Not permitted |
| **Public outdoor space** | Rule of 6 | Rule of 6 | Rule of 6 |
